# Improving mortality prediction in critically ill cancer patients with a multidimensional machine learning model

**DOI:** 10.64898/2026.02.02.26345349

**Authors:** Víctor H Nieto, Adriana C Aya, Andrés F. Cardona, Edwin Pulido, Heidy Trujillo, Natalia Sánchez, Daniel Molano, Nicolle Wagner-Gutiérrez, Oscar Arrieta, Christian Rolfo, Giovanni Nigita, Joseph Nates

## Abstract

**Background:** Prognostic assessment in critically ill patients with cancer remains challenging, as conventional ICU severity scores often perform suboptimally in this population. Machine learning (ML) approaches may improve outcome prediction by integrating acute physiology, organ dysfunction, and oncologic variables. We aimed to develop and validate ML-based models to predict ICU mortality and 30-day survival in critically ill cancer patients.

**Methods:** We conducted a retrospective cohort study including 997 critically ill cancer patients admitted to the ICU. Forty-eight demographic, oncologic, physiological, laboratory, and therapeutic variables collected at ICU admission were used to train and validate ML models. Eight algorithms were evaluated using stratified cross-validation with feature selection and hyperparameter optimization. Model performance was assessed using discrimination, calibration, and classification metrics. Model interpretability was explored using Shapley additive explanations (SHAP).

**Results:** CatBoost achieved the best performance for ICU mortality prediction (AUROC 0.96), showing excellent discrimination and calibration, and outperforming other ML models. Prediction of 30-day survival was less accurate (best AUROC 0.75), reflecting the influence of post-ICU factors not captured at admission. Key predictors of ICU mortality included severity of organ dysfunction, therapeutic objectives, vasopressor and methylene blue use, SAPS III score, lactate, platelet count, and blood urea nitrogen. For 30-day survival, baseline physiological status, admission type, SAPS III, lactate, creatinine, age, and body mass index were most relevant. SHAP analysis demonstrated that acute physiology and organ dysfunction, rather than cancer diagnosis alone, primarily drove short-term outcomes.

**Conclusions:** ML-based models, particularly CatBoost, outperformed traditional prognostic tools for predicting ICU mortality in critically ill cancer patients. Cancer was not an independent predictor of short-term mortality; outcomes were primarily driven by pre-ICU conditions, acute physiology, and severity of organ dysfunction. External validation is needed to confirm generalizability and support future integration of ML-based prediction tools into clinical decision-making in oncologic critical care.

## Introduction

Critically ill patients with cancer are increasingly admitted to intensive care units (ICUs) as advances in oncologic therapies have improved survival and expanded indications for aggressive supportive care(1–4). Despite these advances, this population remains particularly vulnerable, with higher risks of infection, organ dysfunction, and short-term mortality compared with non-oncologic ICU patients(5– 7). Accurate prognostic assessment is therefore essential to guide clinical decision-making, allocation of resources, and discussions regarding goals of care(6).

Traditional ICU severity scores, including APACHE, SOFA, and SAPS, are widely used to estimate prognosis; however, their performance in patients with cancer is limited. Validation studies have shown heterogeneous and often inconsistent results, with frequent underestimation or overestimation of mortality in oncologic populations(8,9). Even prognostic tools specifically developed for patients with cancer have demonstrated variable accuracy across clinical contexts, tumor types, and stages of disease, underscoring the complexity of integrating malignancy-related factors with acute critical illness(8,10,11).

Machine learning (ML) approaches have recently shown promising results in critical care and oncology by integrating multidimensional data and modeling nonlinear relationships. Several ML-based models have improved outcome prediction in general ICU populations and selected oncologic scenarios; however, evidence in critically ill cancer patients remains scarce and largely limited to disease-specific or narrowly defined cohorts(12–17).

Improving prognostic prediction in this population is crucial to reduce bias, support individualized decision-making, and ultimately improve patient outcomes. This study aims to develop and validate ML-based models to predict mortality in critically ill cancer patients admitted to the ICU.

## Methods

### Study design and participants

The dataset used to develop the predictive models was derived from the ambispective EVA (Evidence–Verification–Analysis) cohort. This study was conducted at a high-complexity comprehensive oncology hospital in Bogotá, Colombia, which serves as a national referral center for patients with solid and hematologic malignancies requiring specialized critical care. The study protocol was approved by the local Research and Ethics Committee (Cayre Approval No. 164 of 2022), and written informed consent was obtained from all participants or their legal representatives. Between July 2022 and July 2025, adult patients (≥18 years) with confirmed or suspected malignant neoplasms admitted to the intensive care unit (ICU) for more than 24 hours were eligible for inclusion. The patient selection process is illustrated in Figure 1A. The overall modeling work following, including data preprocessing, model training, validation, and performance evaluation is shown in Figure 1B. The final cohort comprised 997 critically ill cancer patients; 861 were included in the 30-day survival analysis.

### Outcomes and definitions

The primary outcomes were ICU mortality and 30-day survival, both analyzed as binary variables. Candidate predictors were selected based on prior literature identifying prognostic factors in critically ill patients, including those with cancer. A total of 48 variables collected at ICU admission were included. These comprised demographic characteristics (age, sex, body mass index), oncologic variables (primary tumor site, cancer status for solid and hematologic malignancies, performance status, presence of driver mutations, and recent oncologic treatments), physiological and laboratory parameters measured within ±6 hours of ICU admission (including hematologic, renal, metabolic, and coagulation markers), therapeutic interventions (vasopressor use, methylene blue, renal replacement therapy, and mechanical ventilation), and clinical context variables (type of admission, ICU readmission, presence of infection or shock, delirium assessed by CAM-ICU, and severity scores including SAPS III, APACHE II, and SOFA).

### Development of machine learning models

Patients were randomly split into training (70%) and validation (30%) sets. Model robustness and generalizability were ensured using ten-fold stratified cross-validation within the training dataset. Missing continuous variables were imputed within each fold to prevent information leakage, and all features were standardized using z-score normalization. Feature selection was performed using least absolute shrinkage and selection operator (LASSO) regression within each fold to account for collinearity and reduce overfitting. The selected features were then consistently applied across all algorithms.

Two independent prediction models were developed for ICU mortality and 30-day survival using eight machine learning algorithms: Logistic Regression, Random Forest, Extra Trees, Gradient Boosting, AdaBoost, LightGBM, XGBoost, and CatBoost. Hyperparameter optimization was performed within each cross-validation fold. Class imbalance was addressed when applicable, including the use of scale_pos_weight for XGBoost. Model performance was evaluated on the held-out validation sets using discrimination, calibration, and classification metrics, including the area under the receiver operating characteristic curve (AUROC), accuracy, precision, recall, F1-score, specificity, and confusion matrices. Performance metrics were aggregated across folds and visualized to enable comparison across models.

### Model interpretability

To enhance interpretability of the best-performing models, Shapley additive explanation (SHAP) values were computed. SHAP analysis enabled both global and patient-level assessment of feature contributions, allowing visualization of feature importance, directionality of effects, and individual risk drivers through summary, dependence, and decision plots. This approach facilitated transparent interpretation of complex model outputs in clinically meaningful terms.

### Statistical analysis

Continuous variables are reported as medians with interquartile ranges, and categorical variables as counts and percentages. Comparisons between training and validation datasets were performed using the Wilcoxon rank-sum test for continuous variables and Fisher’s exact test for categorical variables. All statistical analyses and model development were conducted using Python 3.11 with open-source libraries, including Pandas and NumPy for data processing, scikit-learn for machine learning, SciPy for statistical testing, and Matplotlib and Seaborn for visualization. A two-sided p-value <0.05 was considered statistically significant. The analytical workflow was designed to ensure reproducibility and transparency. The source code is available upon request or via a public GitHub repository:*(https://github.com/adri1207/Cancer-icu-prediction.git)*.

## Results

### Baseline characteristics

Baseline characteristics provide clinical context in which the machine learning (ML) models were developed and validated. A total of 997 critically ill cancer patients were included in the ICU mortality model, and a subset of 861 patients was analyzed for 30-day survival. Each cohort was randomly divided into training and validation sets (mortality: 697/300; survival: 602/259). Baseline characteristics are summarized in Tables 1 and 2.

In the ICU mortality cohort, the median age was 62 years (IQR, 48–71), and 50.6% of patients were female. The median APACHE II score at admission was 11 (IQR, 8–15), and the median 24-hour SOFA score was 3 (IQR, 2–4.6). Minor differences between the training and validation sets were observed for the presence of shock (p = 0.0336) and renal replacement therapy (p = 0.0099), while all other variables were well balanced.

In the 30-day survival cohort, baseline age and severity scores were comparable between the training and validation sets (APACHE II: 11 [IQR, 8–14]; SOFA: 3 [IQR, 2–4]), and 52% of patients had solid tumors. Statistically significant differences were noted for the PaO_2_/FiO_2_ ratio, partial thromboplastin time, and APACHE II score (all p < 0.05), although overall clinical characteristics remained largely comparable. Taken together, both cohorts demonstrated adequate balance between training and validation subsets, supporting the appropriateness of the random data split for model development.

### Development and assessment of ML models

Among the evaluated algorithms, CatBoost achieved the best performance for ICU mortality prediction, with an AUROC of 0.965, accuracy of 92.5%, F1-score of 0.967, and recall of 94.5%, indicating excellent discrimination and sensitivity. LightGBM showed comparable performance (AUROC 0.960), whereas AdaBoost demonstrated the lowest performance for this outcome (AUROC 0.926), although discrimination remained acceptable. Performance metrics for all models are shown in Figure 2A.

Prediction of 30-day survival was more challenging. CatBoost demonstrated the most balanced overall performance, with an AUROC of 0.754, accuracy of 81.2%, F1-score of 0.392, recall of 29.0%, and precision of 39.2%. Although Extra Trees and Random Forest achieved slightly higher AUROC values (0.765 and 0.761, respectively), CatBoost provided a better balance between sensitivity and precision. AdaBoost again showed the lowest performance (AUROC 0.694), with limited sensitivity for identifying survivors. Comparative model performance for 30-day survival is summarized in Figure 2B.

### Key predictors and model interpretability

Feature importance analysis identified distinct patterns for ICU mortality and 30-day survival prediction. For ICU mortality, ventilatory support emerged as the most influential predictor, followed by therapeutic objectives, methylene blue use, vasopressor support, ICU length of stay, and admission type. Key laboratory and severity markers—including platelet count, blood urea nitrogen, SAPS III score, and lactate level—also contributed substantially to model predictions (Figure 3A). In the 30-day survival model, SAPS III was the most important predictor, followed by direct bilirubin, body mass index, admission type, lactate, sodium, age, and other baseline clinical variables. In contrast to ICU mortality, cancer-related characteristics and pre-ICU physiological status were more influential than acute ICU interventions (Figure 3B).

To enhance interpretability, SHAP analysis was applied to the best-performing models. SHAP summary plots (Figure 4A and 4B) illustrate the global importance of features and the direction of their effects on predicted outcomes. For ICU mortality, the strongest contributions were related to disease stage, therapeutic objectives, organ dysfunction, and acute ICU support. For 30-day survival, predictions were primarily driven by baseline patient characteristics and physiological reserve. SHAP dependence plots highlighted nonlinear relationships and interactions between variables, such as the modifying effect of vasopressor use or disease stage on the association between lactate levels and mortality risk. SHAP decision plots (Figure 4C and 4D) provided patient-level explanations, demonstrating how individual features contributed cumulatively to final risk predictions. Overall, SHAP analysis reinforced the clinical relevance of organ dysfunction, baseline condition, and treatment intensity, while supporting transparent and individualized risk stratification.

## Discussion

In this study, we developed and validated machine learning (ML) models to predict ICU mortality and 30-day survival in a large cohort of critically ill cancer patients. Among the evaluated algorithms, CatBoost demonstrated excellent discrimination for ICU mortality and the most balanced performance for 30-day survival. The models consistently showed that short-term outcomes were primarily driven by acute physiology, severity of organ dysfunction, and treatment intensity rather than by cancer diagnosis alone.

These findings highlight the complex interaction between baseline patient condition, acute critical illness, and therapeutic context in determining outcomes in oncologic ICU patients. The predominance of variables related to cardiovascular instability, tissue hypoperfusion, and organ failure such as vasopressor and methylene blue use, SAPS III score, lactate, platelet count, and renal function suggests that short-term mortality in this population is largely driven by the severity of acute physiological derangements superimposed on the underlying oncologic condition, rather than by cancer diagnosis alone(18,19). Accurate early risk stratification may therefore support more timely and tailored interventions, including not only the initiation but also the appropriate intensity of organ support and therapeutic escalation. Moreover, the ability to estimate risk before ICU admission could facilitate earlier referral and admission, potentially improving outcomes by avoiding delayed access to advanced critical care(20).

In contrast, 30-day survival was more strongly influenced by baseline physiological reserve and chronic patient factors, including admission type, nutritional status, bilirubin, creatinine, and age. These findings indicate that longer-term outcomes are partly determined by pre-existing vulnerabilities and organ dysfunction present at ICU admission, many of which may be less modifiable in the acute setting(21,22). The lower predictive performance observed for 30-day survival likely reflects the contribution of post-ICU factors such as tumor progression, oncologic treatment decisions after discharge, and limitations in therapeutic effort that are not captured by admission-based models and inherently constrain longer-term prediction.

Previous prognostic models in oncology populations have largely relied on traditional regression-based approaches. Karaboyun et al. proposed a mortality prediction model for advanced-stage cancer patients requiring unplanned hospitalization during systemic therapy; however, this model was not designed for ICU patients and lacked external validation(10). Similarly, a systematic review by Cabrera Losada et al. demonstrated that conventional ICU severity scores, including APACHE II, SAPS III, and SOFA, frequently under- or overestimated mortality in patients with cancer, underscoring the limitations of traditional tools in this population(8). In the general critically ill population, ML models have shown incremental improvements over standard severity scores. Nikravangolsefid et al. reported modest but consistent improvements in mortality prediction using machine-learning approaches, particularly logistic regression and eXtreme Gradient Boosting, in large cohorts of patients with sepsis (12), Likewise, Iwase et al. demonstrated high predictive accuracy using random forest models in large, unselected ICU populations, including a cohort of over 12,000 patients, and identified lactate dehydrogenase (LDH) as a novel prognostic biomarker (13).

Within oncologic ICUs, machine-learning applications remain limited, disease-specific, and fragmented in nature. Studies focusing on selected conditions—such as lung cancer, hyperkalemia, or immunotherapy-related toxicity—have demonstrated promising predictive performance but lack generalizability across heterogeneous cancer ICU populations. For example, Huang et al. developed several ML algorithms to predict in-hospital mortality in critically ill lung cancer patients, achieving strong discrimination (AUC 0.92) and identifying SOFA score, albumin, and bilirubin as key predictors(14). Similarly, Bu et al. proposed an interpretable ML model for patients with cancer and hyperkalemia, integrating SHAP and LIME to identify urine output and heart rate as novel prognostic variables(15). Other work, such as that by Wang et al., has explored ML-based prediction of immunotherapy-related toxicity(23). Although these studies highlight the potential value of ML in oncologic critical care, their narrow clinical scope limits applicability to broader and more diverse cancer ICU populations.

From a methodological perspective, this study applied a rigorous and reproducible modeling framework with robust data handling and model development practices. Missing data were imputed using median-based and KNN strategies within the cross-validation folds to prevent information leakage and ensure reliable evaluation. Feature selection was performed using LASSO within each fold, accounting for collinearity while preserving clinically relevant predictors and reducing overfitting. Model training incorporated stratified cross-validation, algorithm-specific hyperparameter tuning, and adjustments for class imbalance (e.g., scale_pos_weight for XGBoost). Multiple machine-learning algorithms were evaluated under identical conditions, with performance metrics aggregated across folds, enabling a stable, comparable, and generalizable assessment of predictive performance across models. The integration of SHAP analysis enhanced interpretability by providing both global and patient-level explanations, supporting transparency and clinical relevance without relying on black-box predictions. In other clinical scenarios such as elderly neurocritical care patients and those with acute respiratory failure SHAP has contributed to identifying key risk factors associated with mortality, potentially prompting earlier interventions or the reevaluation of therapeutic goals(24,25).

Several strengths and limitations should be considered. Strengths include the large real-world cohort, the inclusion of both solid and hematologic malignancies, the evaluation of multiple ML approaches, and the use of explainable AI techniques. However, the study was conducted in a single high-complexity oncology ICU, which may limit generalizability. External validation across diverse centers and healthcare systems is therefore essential. In addition, the lack of granular molecular or genomic oncologic data and the observational design introduce potential residual confounding. Future research should focus on global external validation of these models, incorporation of dynamic time-series data, and prospective evaluation of whether ML-guided risk stratification improves clinical decision-making, ICU triage, and patient-centered outcomes.

In conclusion, ML-based models, particularly CatBoost, demonstrated robust performance in accurately predicting ICU mortality among critically ill cancer patients and consistently outperformed traditional prognostic tools. Our findings indicate that short-term outcomes are driven primarily by acute physiologic derangements and organ dysfunction rather than cancer diagnosis alone. These results underscore the potential of ML-based approaches to enhance mortality and survival prediction in oncologic critical care. External validation is warranted to confirm generalizability and to support the future integration of these models into clinical decision-making frameworks.

## Mandatory declarations

Ethics approval and consent to participate: This study was reviewed and approved by the Cayre Ethics Committee (approval No. 164 of 2025), Bogotá, Colombia. All patients provided informed consent and agreed to the use of data and tumor tissue analysis. Authorization was obtained from the Research Ethics Committee (REC) and Privacy Committee to facilitate the retrospective collection of clinical, pathological, and molecular data.

## Conflicting interest statement

Andrés F. Cardona discloses financial research support from Merck Sharp & Dohme, Boehringer Ingelheim, Roche, Bristol-Myers Squib, Foundation Medicine, Roche Diagnostics, Thermo Fisher, Broad Institute, Amgen, Flatiron Health, Teva Pharma, Rochem Biocare, Bayer, INQBox, and The Foundation for Clinical and Applied Cancer Research – FICMAC. Additionally, he was linked to and received honoraria as an advisor and participated in a speakers’ bureau. He provided expert testimony to EISAI, Merck Serono, Janssen Pharmaceutical, Merck Sharp & Dohme, Boehringer Ingelheim, Roche, Bristol-Myers Squibb, Pfizer, Novartis, Celldex Therapeutics, Foundation Medicine, Eli Lilly, Guardant Health, Illumina, and the Foundation for Clinical and Applied Cancer Research (FICMAC). The other authors have no conflicts of interest to declare.

## Data availability statement

The datasets presented in this article are not readily available because the Colombian organic law of data protection limits access to genetic information in an open format. Requests to access the datasets should be directed to the corresponding author, who will release them upon formal request to the Ministry of Health of Colombia, in accordance with the requirements of Law 1581 of 2012, as amended by the paragraph 201811601170851 of 2018.

## Funding statement

This study did not receive any external funding. The study was conducted using the institutional resources of the Luis Carlos Sarmiento Angulo Treatment and Research Cancer Center, which provided the logistical and operational support required to carry out the study.

## Authors’ contributions

VN and AA planned and coordinated the study; AA, EP, and HT reviewed patient records and composed the database; AA, EP, and HT performed all statistical analyses; VN, AA, EP, HT, and NS performed data interpretation. VN, AA, NW, and AC wrote the initial draft of the manuscript. OA, CR, GN, and JN performed critical revisions and final editing of the manuscript. All authors have contributed to the manuscript and approved the submitted version.

## Notes

### Competing Interest Statement

Andres F. Cardona discloses financial research support from Merck Sharp & Dohme, Boehringer Ingelheim, Roche, Bristol Myers Squib, Foundation Medicine, Roche Diagnostics, Thermo Fisher, Broad Institute, Amgen, Flatiron Health, Teva Pharma, Rochem Biocare, Bayer, INQBox, and The Foundation for Clinical and Applied Cancer Research FICMAC. Additionally, he was linked to and received honoraria as an advisor and participated in a speakers' bureau. He provided expert testimony to EISAI, Merck Serono, Janssen Pharmaceutical, Merck Sharp & Dohme, Boehringer Ingelheim, Roche, Bristol Myers Squibb, Pfizer, Novartis, Celldex Therapeutics, Foundation Medicine, Eli Lilly, Guardant Health, Illumina, and the Foundation for Clinical and Applied Cancer Research (FICMAC). The other authors have no conflicts of interest to declare.

