## Supplemental Table 1 Table 2 and figures for "Improving mortality prediction in critically ill cancer patients with a multidimensional machine learning model"

**Figure 1. (A)** Patient flow diagram. EVA Database; ICU, intensive care unit. **(B)** Model development flowchart.

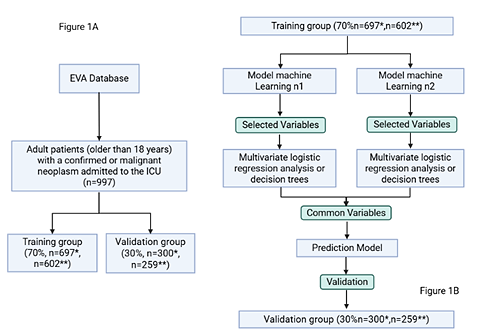

* Mortality dataset ** 30 day-survival dataset

**Table 1.** Baseline patient characteristics mortality model.

| **Patients Mortality** | **All patients**  **N=997** | **Training dataset**  **N=697** | **Validation dataset**  **N=300** | **P-value** |
| --- | --- | --- | --- | --- |
| **Demographic Characteristics** | | | | |
| Age, years | 62 (48-71) | 68 (48-70) | 62 (49-73) | 0.2769 |
| Female | 504 (50.6) | 341(48.9) | 163 (54.3) | 0.1287 |
| BMI | 23.7 (20.7-26.9) | 23.9 (20.6-26.9) | 23.4 (20.8-26.6) | 0.4224 |
| **Oncological history** |  |  |  |  |
| ECOG performance status |  |  |  | 0.1204 |
| *0-2* | 848 (85.1) | 590 (84.7) | 258 (86) |  |
| *3-5* | 149 (14.9) | 107 (15.3) | 42 (14) |  |
| Origin of the tumor |  |  |  | 0.5535 |
| *Solid tumor* | 787 (78.9) | 554 (79.5) | 233 (77.7) |  |
| *Hematologic tumor* | 210 (21.1) | 143 (20.5) | 67 (22.3) |  |
| Staging/risk* |  |  |  | 0.7222 |
| *Advanced* | 305 (30.6) | 212 (30.4) | 93 (31) |  |
| *Early* | 295 (29.6) | 209 (30) | 86 (28.7) |  |
| No data | 397 (39.7) | 276 (39.6) | 121 (40.3) |  |
| Sequencing/molecular study or FISH positive** | 119 (11.9) | 84 (12) | 35 (11.7) | 0.2747 |
| **Physiological parameters** | | | | |
| Lactate, mmol/L | 1.7 (1.2-2.4) | 1.6 (1.2-2.4) | 1.7 (1.3-2.5) | 0.2046 |
| Excess base | -2.9 (-5.0-0.9) | -2.8 (-4.9-0.9) | -3.0 (-5.4-1.0) | 0.3117 |
| Bicarbonate, mmol/L | 20.9 (18.8-22.6) | 21.0 (18.9-22.6) | 20.8 (18.4-22.5) | 0.4807 |
| PaFi ratio | 258 (214-303.3) | 260.4 (215.2-303.3) | 253.7 (204.9-303.5) | 0.2437 |
| Absolute leukocytes, mm³ | 10465 (6392.5-14790) | 10510 (6245-14815) | 10325 (7027.5-14705) | 0.7049 |
| Absolute neutrophils, mcL | 8390 (4450-12507.5) | 8440 (4360-12517.5) | 8255 (4797.5-12377.5) | 0.9942 |
| Absolute lymphocytes, µL | 805.0 (450.0-1367.5) | 780(452.5-1317.5) | 870 (450-1430) | 0.3644 |
| Platelet, µL | 231000(131250-318000) | 227000 (132000-317000) | 243500 (130750-330000) | 0.3681 |
| Hemoglobin, g/dL | 10.8 (8.9-12.9) | 10.8 (8.9-13.0) | 10.8 (9.2-12.6) | 0.8653 |
| Creatinine, mg/dL | 0.8 (0.6-1.1) | 0.8 (0.6-1.1) | 0.8 (0.6-1.1) | 0.2757 |
| BUN, mg/dL | 17.1 (13.2-23.8) | 16.7 (13.0-23.1) | 18.1 (13.5-24.7) | 0.1052 |
| Potassium, mmol/L | 4.2 (3.8-4.5) | 4.2 (3.9-4.5) | 4.1 (3.8-4.6) | 0.4765 |
| Sodium, mmol/L | 137.8 (135.6-140.3) | 137.7 (135.4-140.2) | 138 (135.9-140.5) | 0.2123 |
| Calcium, mg/dL | 8.5 (8-8.8) | 8.5 (8-8.8) | 8.5 (8-8.8) | 0.8682 |
| aPTT, s | 30.3 (28.5-32.8) | 30.5 (28.7-33) | 30.1 (28.2-32.5) | 0.2776 |
| PT, s | 13.6 (12.7-15.5) | 13.6 (12.7-15.5) | 13.7 (12.6-15.4) | 0.4289 |
| Total bilirubin, mg/dL | 0.6 (0.4-1.3) | 0.6 (0.4-1.3) | 0.6 (0.4-1.2) | 0.4577 |
| Direct bilirubin, mg/dL | 0.3 (0.2-0.8) | 0.3 (0.2-0.9) | 0.3 (0.2-0.8) | 0.9839 |
| Indirect bilirubin, mg/dL | 0.3 (0.2-0.4) | 0.3 (0.2-0.4) | 0.3 (0.2-0.4) | 0.2823 |
| ALT, U/L | 42.5 (27.8-69.7) | 41.4 (27.7-69.1) | 43.6 (28.2-72.4) | 0.4686 |
| AST, U/L | 42.4 (31.5-65.3) | 42.1 (30.4-67.4) | 44.0 (32.8-63.6) | 0.3415 |
| **Clinical assessments and ICU admission context** | | | | |
| APACHE II | 11 (8-15) | 11 (8-15) | 12 (8-15) | 0.5442 |
| SAPS III | 43 (32-58) | 43 (31-58) | 45 (34-57) | 0.3135 |
| SOFA | 3 (2-4.6) | 3 (2-4.8) | 3 (2-4) | 0.9631 |
| Type of admission |  |  |  | 0.7268 |
| *Medical* | 575 (57.5) | 399 (57.3) | 176 (58.7) |  |
| *Elective/emergency surgery* | 421 (42.3) | 297 (42.7) | 124 (41.3) |  |
| Therapeutic objective |  |  |  | 0.2798 |
| *Therapeutic* | 895 (89.8) | 632 (90.7) | 263 (87.7) |  |
| *Palliative* | 79 (7.9) | 49 (7) | 30 (10) |  |
| *Not established* | 23 (2.3) | 16 (2.3) | 7 (2.3) |  |
| Sepsis | 288 (28.9) | 206 (29.6) | 82 (27.3) | 0.4938 |
| Shock*** | 331 (33.2) | 246 (35.3) | 85 (28.3) | 0.0336 |
| Delirium | 164 (16.4) | 117 (16.8) | 47 (15.7) | 0.7099 |
| **Therapeutic interventions and support measures** | | | | |
| Ventilatory support**** | 302 (30.3) | 210 (30.1) | 92 (30.7) | 0.8807 |
| Painkillers use | 732 (73.4) | 515 (73.9) | 217 (72.3) | 0.6393 |
| Vasopressors use | 407 (40.8) | 289 (41.5) | 118 (39.3) | 0.5742 |
| Sedatives use | 253 (25.4) | 176 (25.3) | 77 (25.7) | 0.9368 |
| Methylene blue use | 61 (6.1) | 20 (6.7) | 41 (5.9) | 0.6661 |
| Oncological treatment***** | 68 (6.8) | 51 (7.3) | 17 (5.7) | 0.4116 |
| Renal support****** | 43 (4.3) | 22 (3.2) | 21 (7) | 0.0099 |
| **Other characteristics** | | | | |
| LOS, days | 3 (2-5) | 3.0 (2-4) | 3 (2-5) | 0.7691 |
| ICU readmission |  |  |  | 0.0961 |
| *1 admission* | 443 (52.6) | 297 (50.5) | 146 (57.3) |  |
| *2 admissions* | 240 (28.5) | 180 (30.6) | 60 (3.5) |  |
| *>3 admissions* | 160 (19) | 111 (18.9) | 49 (19.2) |  |

Continuous variables are presented as medians and interquartile ranges (IQRs) and were compared using the Wilcoxon rank sum test. Categorical variables are presented as counts and percentages and were compared using Fisher’s exact test. IQR interquartile range, Pressure of Arterial Oxygen / Fraction of Inspired Oxygen, ALT alanine aminotransferase, AST aspartate aminotransferase, APTT activated partial thromboplastin time, PT partial thromboplastin time, APACHE II Acute Physiology and Chronic Health Evaluation, APS III Acute Physiology Score III, SOFA Sequential Organ Failure Assessment, length of stay in the ICU

*Staging in solid tumors, lymphomas and myelomas early or advanced, Risk in leukemias high or low **Solid tumor sequencing study, molecular study in leukemias or FISH in myelomas or lymphomas positive *** If the patient presented with any of the following types of shock: Hypovolemic, Obstructive, Distributive, Cardiogenic **** If the patient required any ventilatory support non-invasive mechanical ventilation, high-flow oxygen therapy, or invasive mechanical ventilation ***** If the patient received any type of oncological treatment, such as immunotherapy, chemotherapy, targeted therapy, hormonal therapy, or radiotherapy ****** If the patient required renal replacement therapy, such as hemodialysis or peritoneal dialysis.

**Table 2.** Baseline patient characteristics 30-day survival model.

| **Patients 30-day survival** | **All patients**  **N=861** | **Training dataset**  **N=602** | **Validation dataset**  **N=259** | **P-value** |
| --- | --- | --- | --- | --- |
| **Demographic characteristics** | | | | |
| Age, years | 62 (48-71) | 61 (48-71.8) | 63 (50-71) | 0.4199 |
| Female | 442 (51.3) | 311 (51.7) | 131 (50.6) | 0.8236 |
| BMI | 23.7 (20.8-26.9) | 23.8 (20.9-27) | 23.5 (20.7-26.5) | 0.3598 |
| **Oncological history** |  |  |  |  |
| ECOG performance status |  |  |  | 0.9801 |
| *0-2* | 743 (86.2) | 519 (86.2) | 224 (86.5) |  |
| *3-5* | 118 (13.8) | 83 (13.8) | 35 (13.5) |  |
| Origin of the tumor |  |  |  | 0.2933 |
| *Solid tumor* | 701 (81.4) | 496 (82.4) | 205 (79.2) |  |
| *Hematologic tumor* | 160 (18.6) | 106 (17.6) | 54 (20.8) |  |
| Staging/risk* |  |  |  | 0.2515 |
| *Early* | 267 (31) | 195 (32.4) | 72 (27.8) |  |
| *Advanced* | 261 (30.3) | 178 (29.6) | 83 (32) |  |
| No data | 333 (38.7) | 229 (38) | 104 (40.2) |  |
| Sequencing/molecular study positive** | 99 (11.5) | 75 (12.5) | 24 (9.3) | 0.2476 |
| **Physiological parameters** | | | | |
| Lactate, mmol/L | 1.6 (1.2-2.3) | 1.6 (1.2-2.3) | 1.7 (1.3-2.3) | 0.7348 |
| Excess base | -2.8 (-4.7-0.9) | -2.8 (-4.7-0.9) | -2.8 (-4.6-1) | 0.8743 |
| Bicarbonate, mmol/L | 21 (19-22.6) | 21 (19 -22.6) | 21 (19-22.5) | 0.4934 |
| PaFi ratio | 263.6 (223.6-306.8) | 261.4 (221.1-303.2) | 270 (226.9-319.3) | 0.0478 |
| Absolute leukocytes, mm³ | 10675 (6905 -14840) | 10400 (6925-14850) | 11150 (6840-14750) | 0.4502 |
| Absolute neutrophils, mcL | 8635 (4910-12585) | 8480 (4850-12675) | 8930 (5180 -12430) | 0.5675 |
| Absolute lymphocytes, µL | 870.0 (500-1395) | 800 (490-1380) | 855 (500-1382.5) | 0.7294 |
| Platelet, µL | 244000 (159750-327500) | 245000 (159000-320000) | 241000 (161000-334000) | 0.843 |
| Hemoglobin, g/dL | 11 (9.1-13) | 11.1 (9.2-13.1) | 10.8 (9-12.9) | 0.345 |
| Creatinine, mg/dL | 0.8 (0.6-1) | 0.8 (0.6-1) | 0.8 (0.6-1.1) | 0.9627 |
| BUN, mg/dL | 16.7 (13-22) | 16.7 (12.7-21.7) | 16.6 (13.7-23.6) | 0.1781 |
| Potassium, mmol/L | 4.2 (3.9-4.5) | 4.2 (3.9-4.5) | 4.2 (3.8-4.5) | 0.9753 |
| Sodium, mmol/L | 138 (135.6-140.3) | 138 (135.6-140.3) | 138 (135.6-140.4) | 0.9311 |
| Calcium, mg/dL | 8.5 (8.1-8.9) | 8.6 (8.1-8.9) | 8.5 (8-8.8) | 0.0884 |
| aPTT, s | 30.2 (28.4-32.6) | 30 (28.2-32.2) | 30.9 (28.8-33) | 0.008 |
| PT, s | 13.6 (12.6-15.4) | 13.5 (12.5-15.2) | 13.8 (12.8-15.7) | 0.0142 |
| Total bilirubin, mg/dL | 0.6 (0.4-1.3) | 0.6 (0.4-1.3) | 0.6 (0.5-1.3) | 0.1942 |
| Direct bilirubin, mg/dL | 0.3 (0.2-0.8) | 0.3 (0.2-0.8) | 0.3 (0.2-0.8) | 0.1016 |
| Indirect bilirubin, mg/dL | 0.3 (0.2-0.4) | 0.3 (0.2-0.4) | 0.3 (0.2-0.4) | 0.2392 |
| ALT, U/L | 42.5 (28.2-69.1) | 43 (28.3-69) | 41.3 (27.9-70) | 0.7944 |
| AST, U/L | 41.3 (30.9-63.6) | 41.6 (31-63.3) | 40.7 (30.3-64.3) | 0.9782 |
| **Clinical assessments and ICU admission context** | | | | |
| APACHE II | 11 (8 -14) | 11 (8-14) | 12 (8-15) | 0.0261 |
| APS III | 41 (31-55) | 40 (30-55) | 44 (32-55.2) | 0.0841 |
| SOFA | 3 (2-4) | 3 (2-4) | 3 (1.2-4) | 0.9765 |
| Type of admission |  |  |  | 0.3331 |
| *Medical* | 447 (52) | 306 (50.8) | 141 (54.7) |  |
| *Elective/emergency surgery* | 413 (48) | 296 (49.2) | 117 (45.3) |  |
| Therapeutic objective |  |  |  | 0.5 |
| *Therapeutic* | 810 (94.1) | 565 (93.9) | 245 (94.6) |  |
| *Palliative* | 38 (4.4) | 26 (4.3) | 12 (4.6) |  |
| *Not established* | 13 (1.5) | 11 (1.8) | 2 (0.8) |  |
| Sepsis | 201 (23.3) | 131 (21) | 70 (27) | 0.096 |
| Shock*** | 226 (26.2) | 77 (29.7) | 149 (24.8) | 0.1295 |
| Delirium | 128 (14.9) | 81 (13.5) | 47 (18.1) | 0.0942 |
| **Therapeutic interventions and support measures** | | | | |
| Ventilatory support**** | 177 (20.6) | 24 (20.6) | 53 (20.5) | 1 |
| Painkillers use | 602 (69.9) | 426 (70.8) | 176 (68) | 0.4186 |
| Vasopressors use | 189 (31.4) | 96 (37.1) | 285 (33.1) | 0.1144 |
| Sedatives use | 107 (17.8) | 44 (17.0) | 151 (17.5) | 0.8452 |
| Methylene blue use | 9 (1) | 5 (0.8) | 4 (1.5) | 0.4645 |
| Oncological treatment***** | 50 (5.8) | 37 (6.1) | 13 (5) | 0.634 |
| Renal support****** | 24 (2.8) | 13 (2.2) | 11 (4.2) | 0.1125 |
| **Other characteristics** | | | | |
| LOS, days | 3 (2-4) | 3 (2-4) | 2 (2-4) | 0.8758 |
| ICU readmission |  |  |  | 0.9002 |
| *1 admission* | 391 (52.9) | 274 (52.5) | 117 (53.9) |  |
| *2 admissions* | 213 (28.8) | 153 (29.3) | 60 (27.6) |  |
| *>3 admissions* | 135 (18.3) | 95 (18.2) | 40 (18.4) |  |

Continuous variables are presented as medians and interquartile ranges (IQRs) and were compared using the Wilcoxon rank sum test. Categorical variables are presented as counts and percentages and were compared using Fisher’s exact test. IQR interquartile range, Pressure of Arterial Oxygen / Fraction of Inspired Oxygen, ALT alanine aminotransferase, AST aspartate aminotransferase, APTT activated partial thromboplastin time, PT partial thromboplastin time, APACHE II Acute Physiology and Chronic Health Evaluation, APS III Acute Physiology Score III, SOFA Sequential Organ Failure Assessment, length of stay in the ICU

*Staging in solid tumors, lymphomas and myelomas early or advanced, Risk in leukemias high or low **Solid tumor sequencing study, molecular study in leukemias or FISH in myelomas or lymphomas positive *** If the patient presented with any of the following types of shock: Hypovolemic, Obstructive, Distributive, Cardiogenic **** If the patient required any ventilatory support non-invasive mechanical ventilation, high-flow oxygen therapy, or invasive mechanical ventilation ***** If the patient received any type of oncological treatment, such as immunotherapy, chemotherapy, targeted therapy, hormonal therapy, or radiotherapy ****** If the patient required renal replacement therapy, such as hemodialysis or peritoneal dialysis.

**Figure 2. (A)** Comparison of metrics by model and target. **(B)** ROC Curves by model and target.

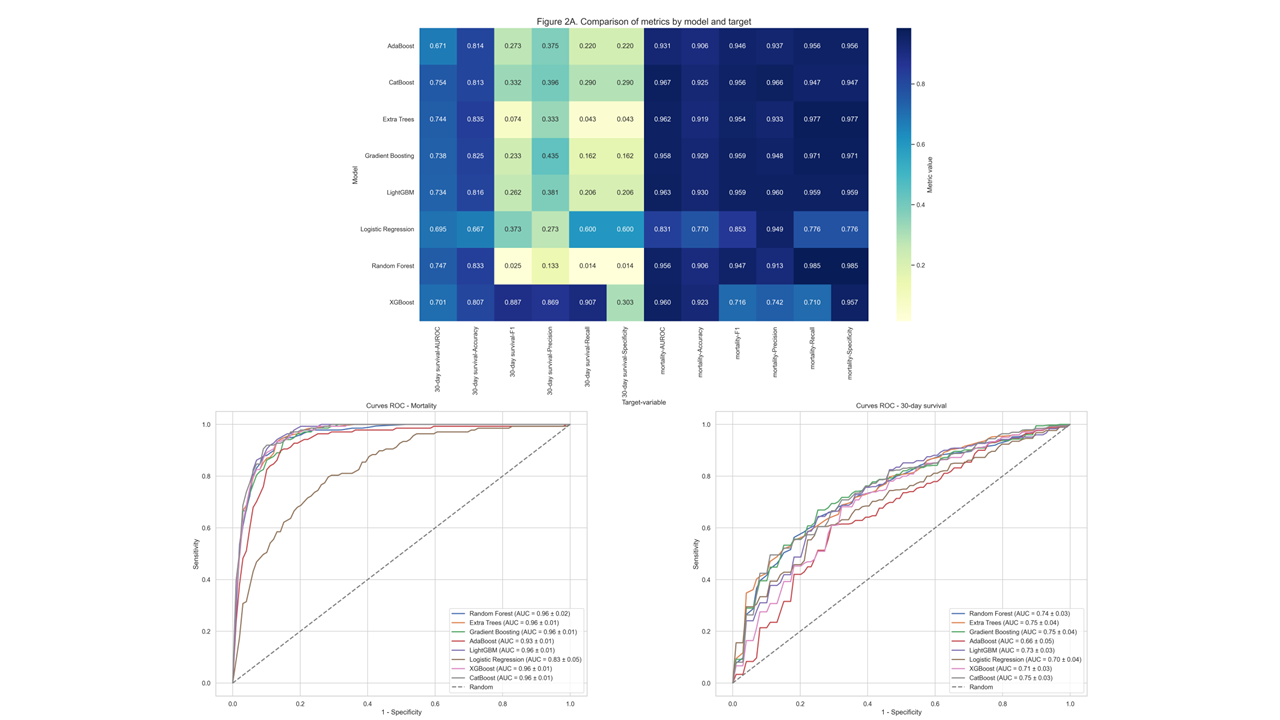

**Figure 3.** Feature importance plots for all variables. **(A)** Feature importance plot of CatBoost mortality. **(B)** Feature importance plot of CatBoost 30-day survival.

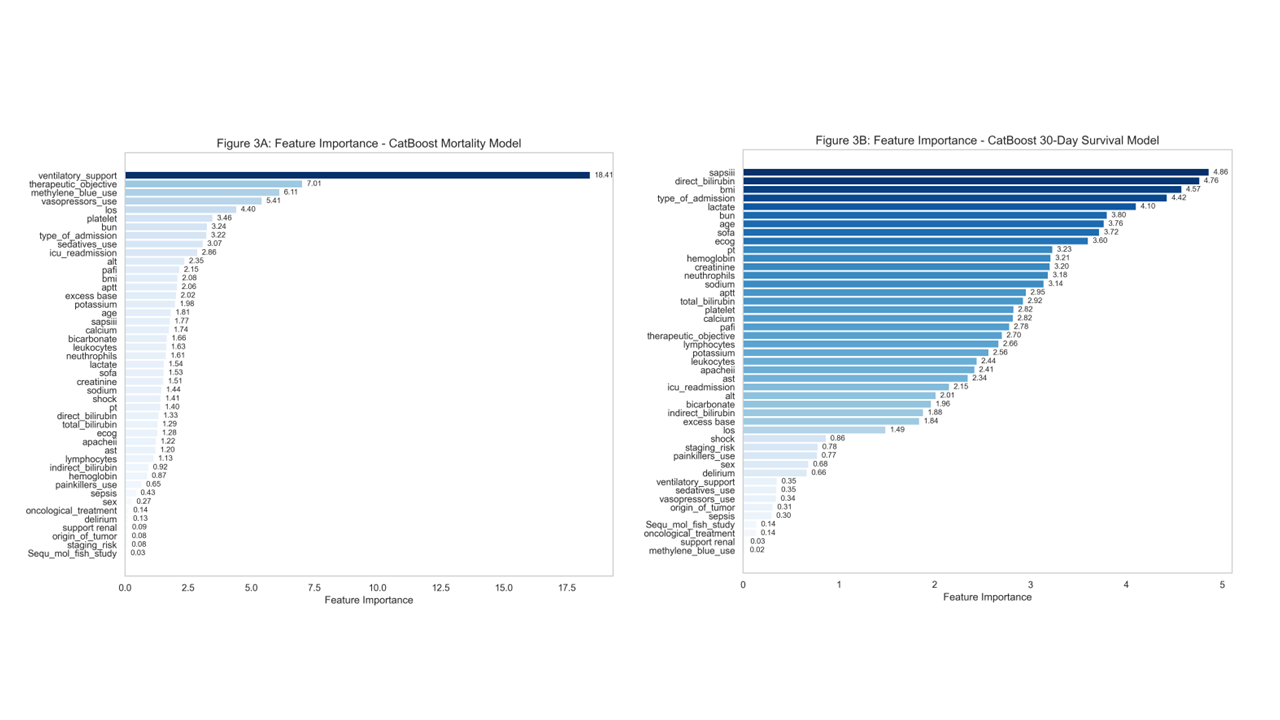

**Figure 4.** SHAP analysis of the final CatBoost for both models. **(A)** SHAP summary plot mortality model. **(B)** SHAP summary plot 30-day-survival model. **(E)** SHAP decision plot mortality model. **(F)** SHAP decision plot 30-day-survival model.

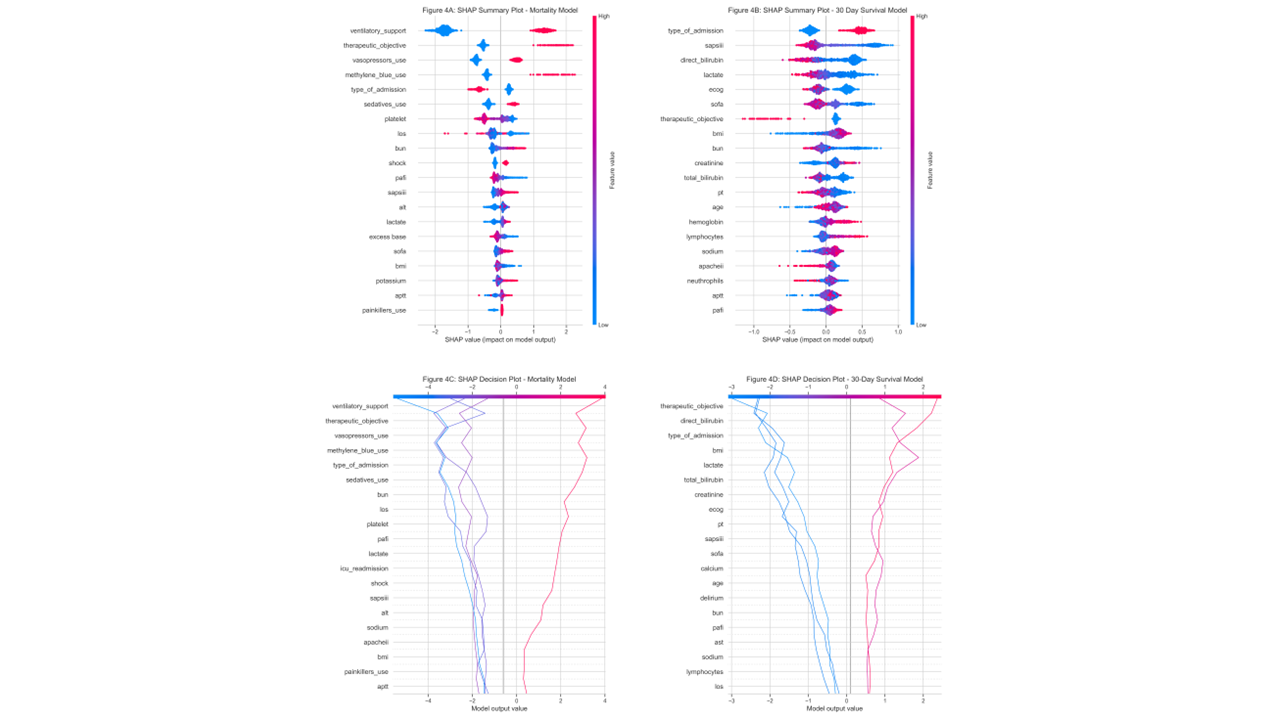
